# Scaling Laws in Healthcare Delivery. A Network Analysis Approach to Outpatient Referral Networks Across the Veterans Health Administration

**DOI:** 10.1101/2024.11.06.24316731

**Authors:** Alon Ben-Ari, Harry L. George, Paul E. Bigeleisen

## Abstract

**BACKGROUND:** We studied clinical referral networks across the Veterans Health Administration. Network attributes, referral counts and timeliness of services rendered were estimated and used to predict outpatient care performance. A scaling law for outpatient referrals was derived, predicting an economy of scale.

**METHODS:** Network topology and operational statistics were estimated and their relationships were studied. A comparative study of multiple networks was undertaken.

**RESULTS:** We observed similarities both in network structure and in the referral patterns. These are invariant of the healthcare system size. A power scaling law for referrals was inferred with an infra-linear exponent.

**CONCLUSION:** Study of referral networks across multiple healthcare systems shows an economy of scale and self-similarity between network attributes that are patient size invariant.

## Introduction

The majority of health care is delivered in the outpatient setting where placing a referral for a consultation and the completion of that consult is a common measure. This entity, called access to care, is a major metric for healthcare systems, impacting revenue, service quality and predicting the patient referral given a patient population. In the Veterans Health Administration (VHA), this has been the focus of two major legislation efforts (*1, 2*). To better study this, we used a data base of referring disciplines, (i.e. referral from cardiology to vascular surgery). We also measured referral completion time attributes and referral counts. Using an operational metric, along with network attributes, we modelled those network behaviors which would provide an improved understanding. This includes the effect of scale on the economy of health care. The question of scaling laws has been studied in many systems, ranging from biology (*3*), urban science (*4, 5*), finance (*6*) and ecology (*7*). Thus, we assumed that using methodologies from those fields to study scaling laws in healthcare delivery would allow us to answer this question. We characterized referral network referrals comprising of 130 healthcare systems (HCS) across the entire VHA during a one year period. Specifically, we describe network attributes and modelled their scaling laws as a function of their corresponding patient base size. We also describe their access to care time as a function of network features showing similarities in referral patterns between healthcare systems which are invariant of size. Finally, we point to future directions of applications that can be built from the observations we make.

## Results

Network construction is detailed in (*8*). Functional clinical and administrative units serve as network nodes. An edge between nodes exists if a referral was made. Median time to referral completion and count of referrals for that period serve as edge attributes. This was accomplished for all 130 systems in the VHA. Analysis (*8*) was on network descriptive statistics of weighted in degree distribution, transitivity, clustering, density and average path (see glossary in supplemental text.) Results are seen in figure 1 and figure 2. The narrow range of values shows similarity between network geometry. Specifically, log versus log plotting of in degree distribution results shows scale free networks that are invariant of the size of the networks (figure 2). We employed spectral distance analysis (*9*) to further study the difference between networks (figure 3). From this we observed size dependent self similarity (figure 3). That is, large net works are closer in their structure. In contrast, smaller health care systems have their own structure which is a scaled version of larger ones. Also we explored similarity of several key systems. Using cardiology referral patterns across all VHA systems (figure 4) shows general agreement (using cosine similarity) across VHA for this referral pattern. Next, we used a predictive model (*8*) for the expected number of referrals given a patient population base (5). Model fitting performance in table 1 shows an infra-linear power law as a scaling law for the expected number of future face to face visits. This model has a better fit that a simple linear modeling. This implies, that with a growing population base, the expected count of referrals will go down, that is, an economy of scale. Finally we explored the relationship between network geometric features and wait times. All measures were transformed to normalized z-scores to mitigate difference of scales. Results in figure 6, show an inverse association between the networks transitivity and their density and timeliness. As expected, healthcare systems serving a larger population have networks of higher connectivity and are able to process consult requests faster. This is the first observation that network topology features are tied to operational features for such a network

**Table 1:** scaling law for expected number of referrals given patient population size. Model fitting statistics forA power-law model suggests a superior fit with an exponent of 0.823.

| model | coeff | p-values | mse | aic | loglike |
| --- | --- | --- | --- | --- | --- |
| ax+b | 0.661304 | 6.481538e-23 | 8.511313e+10 | 2971.189503 | -1483.594752 |
| logy = log(a)+blogx | 0.823589 | 2.467516e-35 | 3.139678e+01 | 76.300543 | -36.150271 |

**Figure 1:**
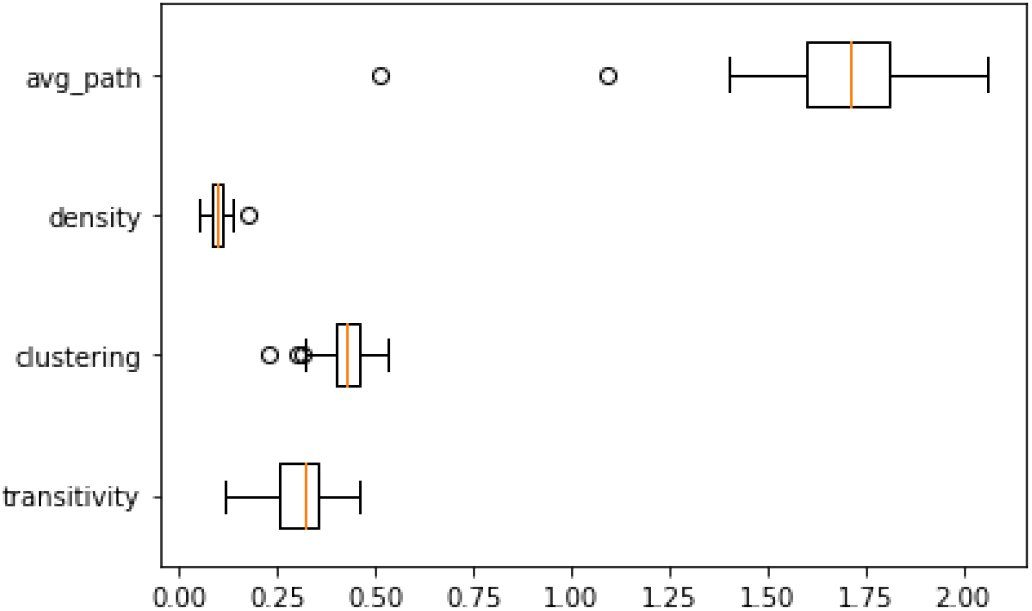
Distribution of some key network measures. Notice tight ranges for transitivity, density, average path and clustering coefficient across VHA’s healthcare system across the US.

**Figure 2:**
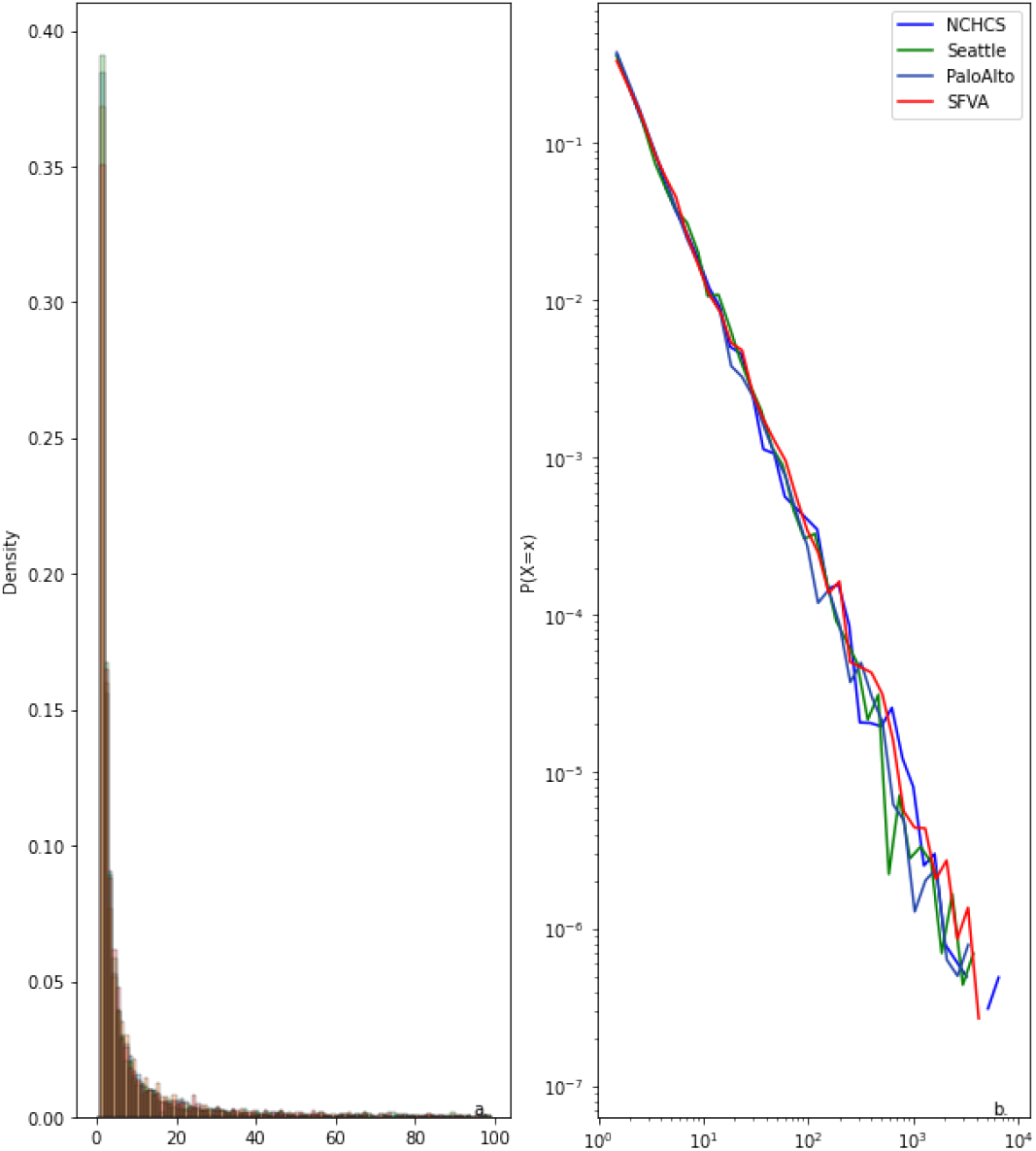
Distribution of some key network measures. Notice tight ranges for transitivity, density, average path and clustering coefficient across VHA’s healthcare system across the US.

**Figure 3:**
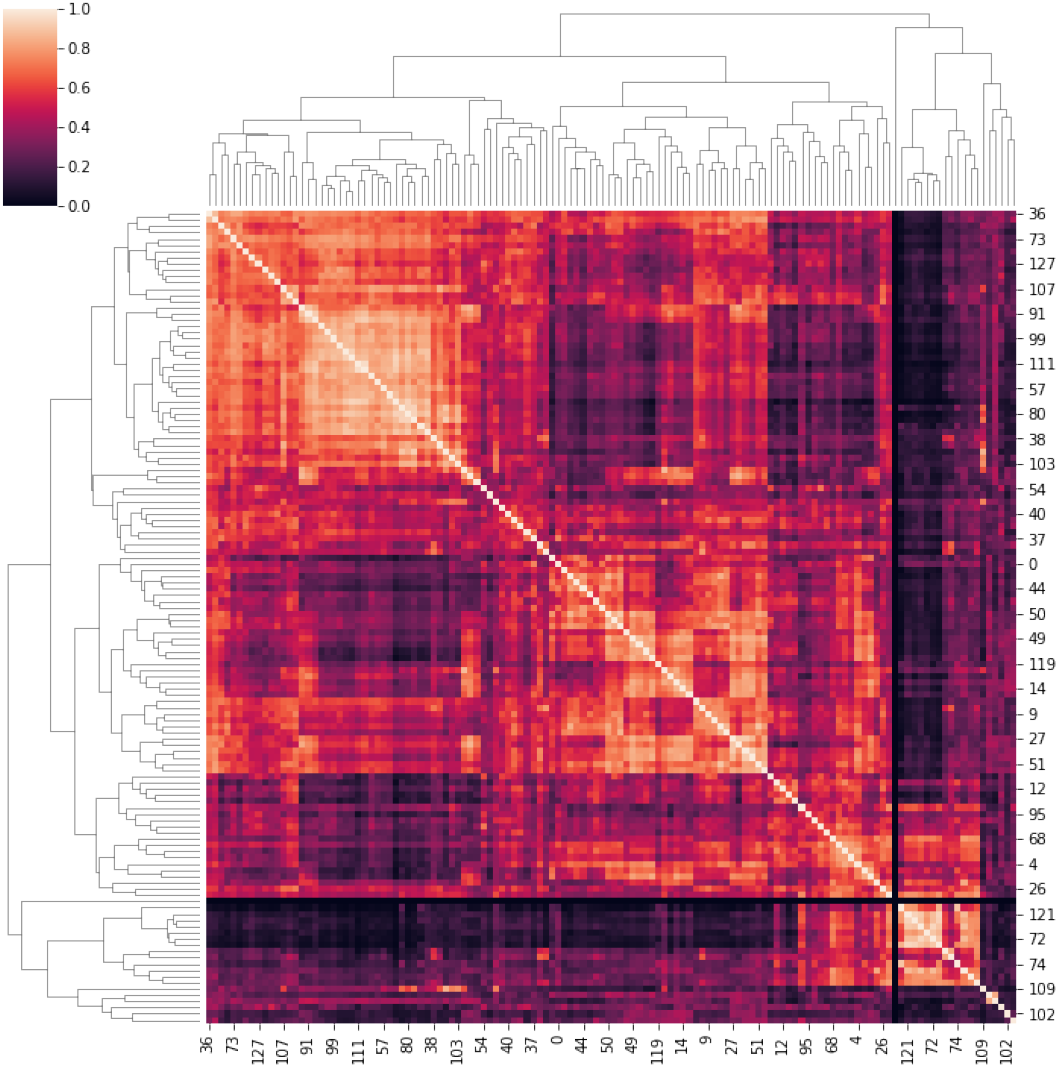
Heatmap of similariy between networks using eigen-methods. Networks were ranked by the catchment area size. The visual suggests greater similarity between networks of similar catchment area. sample.’

**Figure 4:**
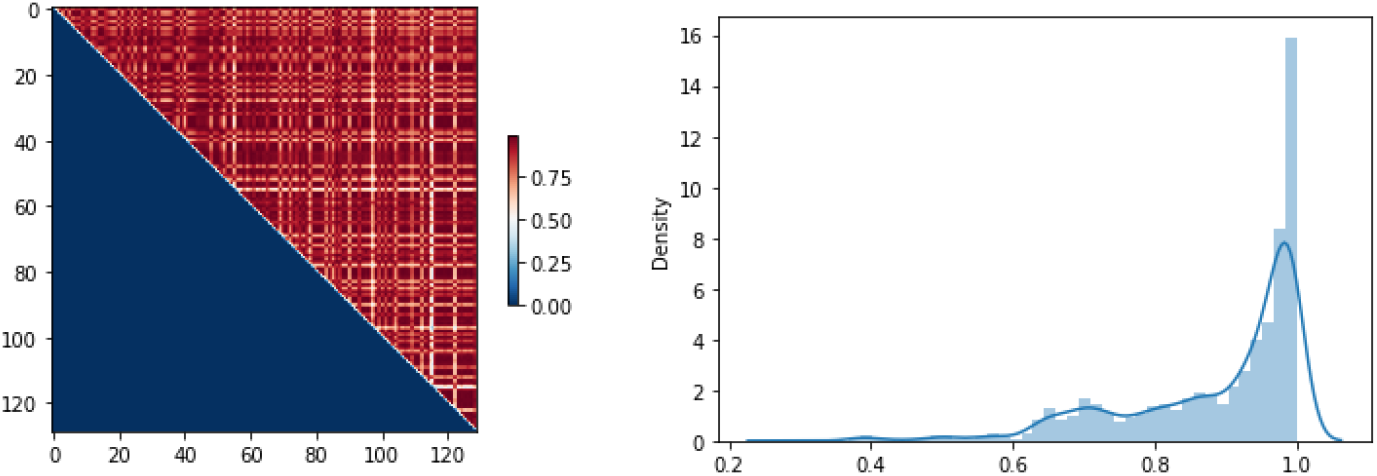
Similarity in pattern of referrals in cardiology across VHA. (left) Upper triangle heatmap of cosine similarity of incoming referrals into cardiology across VHA. (right) Cosine similarity density of incoming referrals for cardiology, across VHA. Median(IQR): 0.95(0.81-0.98

**Figure 5:**
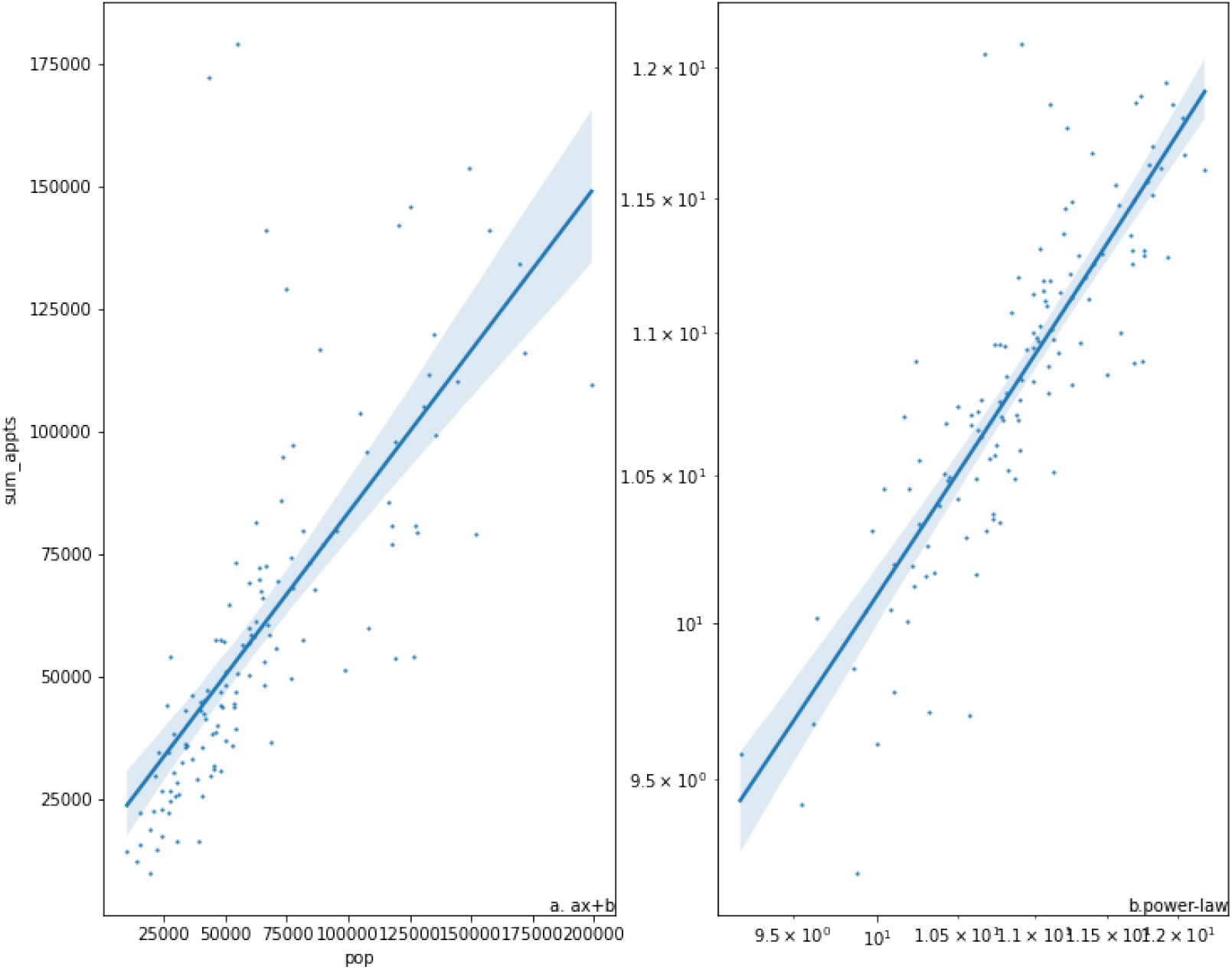
Scaling-law for modeling the change of expected number of referrals given a set population base. a. shows a linear model, b. shows model fitting for a log-log transformation. Statistics are detailed in table 1. suggesting superior fit for a power law model.

**Figure 6:**
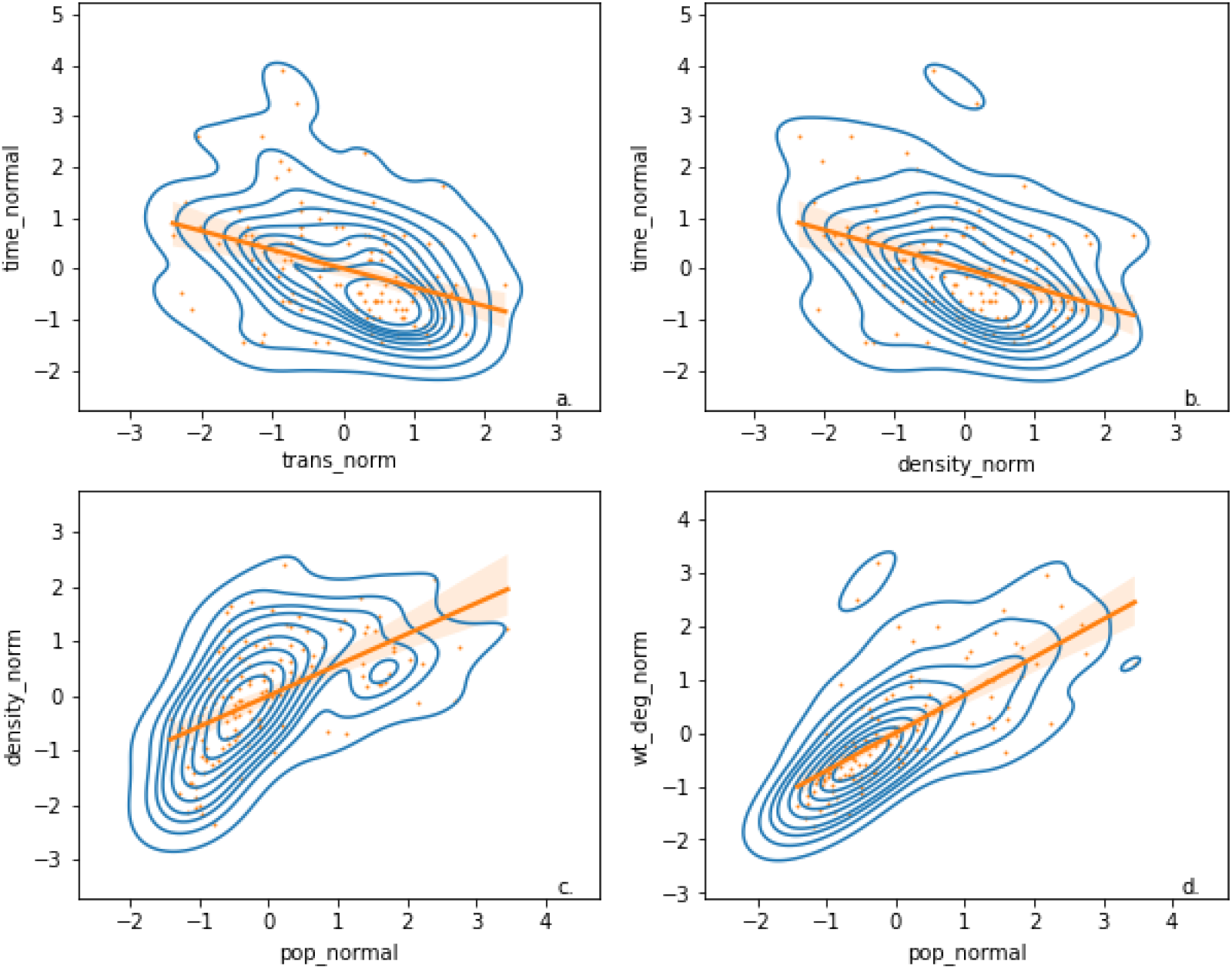
z-scores of density, patient population size and transitivity of networks and its association to operational.. a. Normalized transitivity and normalized time to service b. Normalized density and normalize time to service. c. Association between normalized population size and normalized density. d. z-scores weighted in-degree centrality as a function of normalized population. Suggested linear relationship are plotted.

## Discussion

This is the first study that formally compares multiple outpatient referral networks across one of the largest healthcare systems in the United States. We used a formalism from network analysis from others (*3–7*) and applied it to outpatient healthcare delivery. Our choice of a network model, proved useful and highlighted that underlying the outpatient referral system, is a conserved network structure. This structure holds regardless of the variability of factors, including academic affiliation, different offering of clinical services and geography, and variance in clinical practice. Thus, this model formally and succinctly captures the essence of outpatient healthcare delivery. In essence, thousands of VHA staff members, independently taking care of veterans across the United States, gives rise to a conserved pattern that this study revealed. We showed that the expected number of referrals in a healthcare system follow a power law with an exponent less is than 1, that is infra-linear. This means that as the patient population grows by a certain factor, the number of referrals will scale by less than that factor, that is, an economy of scale. The power law we found is also associated with self similarity or a scale free phenomenon, so that a given referral network is a scaled version of any other healthcare system, which may be larger or smaller, and share the same features. In trying to quantify the differences between the networks, we graphically captured the distance between networks in a heat map (figure 3). This points to an additional observation: Ranking the networks by patient catchment size, we observe that healthcare systems’ referral networks of similar catchment size, have a cosine similarity close to 1.0 between their eigen-values. This underlies the idea of self-similarity between referral networks that serve similar population sizes. Access to care time in the VHA has been the focus of legislation, that is, the Choice ACT (1) and the Mission ACT (2) which allow veterans to receive care outside the VHA. Similarly, access to care in Medicare, Medicaid and the commercial insurance industry impacts patent care quality, including retention of patients and revenue. Our work shows those factors which predict access to care time and predict the patient referral load given a patient population. Referral networks attributes and their narrow variability shows that a generative model can be derived to meet operational needs in predicting and modifying access to care. We formally define a scaling law for referrals in healthcare system

## Conclusion

This work is the beginning of putting healthcare management on a formal foundation with a principled framework to explore access to healthcare and healthcare operations.

## Data Availability

Data is currently NOT available, pending VHA's research office approval.

## Acknowledgments

VINCI, VA INformatics Computing Infrastructure. Office of Research and Development.

## Author contributions

HG SQL development of cohort, PB editorial, analytical. AB manuscript authorship, data analysis, modelling, visualization.

## Supplementary materials

Supplementary Text

Figs. S1 to S3

Tables S1 to S4

References (*7-15*)

Movie S1

Data S1

## Materials and Methods

### 0.0.1 Network construction

VHA is composed of 130 health care systems (HCS). Following Northern California Healthcare System’s (NCHCS) Institutional Review Board’s approval, referral data from VA INformatics and Computing Infrastructure (VINCI) (*10*) were obtained for the period 12/31/2018 through 12/31/2019 for VHA’s systems across the country. Only referrals that were *scheduled and resulted in a visit* were considered for analysis. For administrative purposes, VHA assigns a code for each clinical or admin service known as ’Stop-Code’ which are conserved across all VHA. To model the referral network, we use,’Stop codes’ as the nodes in the network. Directed edges between nodes exist if a referral existed between clinical services (i.e. Stop codes) during the said period. Since *Stop-Codes are conserved across VHA*, networks whose nodes are stop-codes allowed us to compare the different networks from different HCS across the country. Counts of referrals and the median time to referral completion were used as edge attributes. The size of the health care system were defined as the count of their patient base size which were abstracted from VINCI.

### 0.02 Analysis

- Network realization: Referral networks were realized for each healthcare system across VHA using Python’s Networkx package (*11, 12*). d
- Network features: The Average shortest path, clustering, density (fraction of actual edges to possible edges), weighted degree distribution, and transitivity (fraction of triangles in a graph to possible triangles) were computed for all networks. Summary statistics of the normalized degree distribution of the weighted (equivalent to referral counts) networks were computed.
- Distribution of weighted (count of referrals) in-degree:The weighted in-degree density functions were normalized and plotted on a log-log scale. The total number of consults in a healthcare system were computed as a function of the HCS patient size.
- comparing different networks: The network’s adjacency matrix were ranked from largest to smallest by patient enrollee size. We used the spectral distance (*9*)

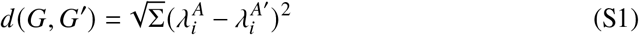

where *G, G*^′^ are the graphs, 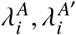 are the eigenvalues of the adjacency matrices of *G, G*^′^ respectively. A matrix whose axes were ranked by size of healthcare systems was used to record the the measure d (*G, G*^′^) the result is reported as a heat map.
- Comparing the outgoing or incoming referrals across all Healthcare system: A vector containing weighted edges of outgoing edges from a specific stop-code were normalized. (Alternatively, for incoming referrals, weighted edges going into a chosen stop code are used). A matrix containing the cosine similarity (*13*) between all possible referral patterns across VHA were computed. Since all vectors had a length of one unit, referral load was mitigated and the only difference would be the angle between the vectors. Technically, since the matrix contained two cosine similarity measures for each pair of healthcare system, the lower triangle of the matrix was removed from analysis. The values of the matrix diagonal were remove too because they would always carry the value of one. A density plot of the values was used to assess the degree of similarity between the referral patterns. A median, Inter Quartile Range (IQR) and a histogram were used to appreciate the degree of similarity.
- Scaling laws for the expectation of count of referrals: We estimated the expected value of counts of referrals as a function of the patient population. A simple linear model and a log-log transformed linear model were fit to the data. We compared the log-likelihood, and AIC to compare the two models. 2) We modeled Expectation of mean access to care time as a function of the density of the network. 3) We studied how transitivity (count of triangles to potential number of triangles in a network) and density (ratio of current edges potential edges) scales with patient population size and its relation to timeliness of access. Fro the purpose of analysis we normalized the values for time, density and transitivity, and obsevre the relationship and variability across VHA.
- Software packages: Numerical analysis were carried using Python’s numpy (*14*), scipy (*15*), networksx (*12*).

## Supplementary Text

## Appendix

### In-degree centrality of a node

In-degree of a node is defined as the number of incoming edges into a node.

### Weighted in-degree centrality

Weighted in degree is the sum of the weights of the in-coming edges.

### Density

The proportion of existing edges to potential edges.

### Transitivity

The proportion of existing triangles to potential triangles.

### Average path

The average number of edges along the shortest path for all possible pairs of nodes in the network.

### Spectrum of a graph/Spectral graph methods

The study of graph properties as they relate the eigenvalues and eigen-vectors of their adjacency matrix, and their Laplacian matrix.

### Eigen-Vector/Eigen-value

In linear algebra, a matrix is associated with a unique eigen-value, eigen-vector pairs, where by the matrix multiplied by the eigen-vector returns the same eigen vector scaled by the eigen-value. The Supplementary Text section can only be used to directly support statements made in the main text e.g. to present more detailed justifications of assumptions, investigate alternative scenarios, provide extended acknowledgements etc. Material in this section cannot claim results or conclusions that weren’t mentioned in the main text. To refer to this section from the main text, just write (Supplementary Text).

